# Ezetimibe and Atherosclerotic Cardiovascular Disease: A Systematic Review and Meta-Analyses

**DOI:** 10.1101/2023.07.29.23293356

**Authors:** Fatemeh Omidi, Maryam Rahmannia, Parsa Mohammadsharifi, Mohammad Javad Nasiri, Tala Sarmastzadeh

## Abstract

**Introduction:** Individuals diagnosed with atherosclerotic cardiovascular disease (ASCVD), particularly those who have experienced acute coronary syndrome (ACS) within the past year, are at a heightened risk of recurrent cardiovascular events. Lowering low-density lipoprotein cholesterol (LDL-C) levels has been proven effective in reducing this risk. However, there is a lack of a comprehensive meta-analysis investigating the LDL-C-lowering effectiveness and coronary atherosclerotic plaque compositions of Ezetimibe. This study aims to address this gap in knowledge.

**Methods:** We conducted a systematic review of randomized controlled trials that evaluated the LDL-C-lowering efficacy and coronary atherosclerotic plaques efficacy of ezetimibe in patients with ASCVD. We searched relevant databases, including MEDLINE, EMBASE, and the Cochrane Central Register of Controlled Trials, for publications from database inception until Jul 2023. Meta-analyses were performed to assess the LDL-C-lowering and coronary atherosclerotic plaques efficacy of ezetimibe in the overall ASCVD population.

**Results:** The meta-analysis included a total of 20 eligible studies. Our findings revealed that combination therapy of ezetimibe with statins resulted in a more substantial absolute reduction in LDL-C compared to statin monotherapy (mean difference of (−14.06 mg/dL; 95% confidence interval [CI] −18.0 to −10.0; p = 0.0001) after 6-12 months of treatment (or at a time point nearest to 6 -12months). Moreover, the subgroups analysis indicates that the intervention measures were effective in reducing the volume of fibro-fatty plaque (FFP) when compared to the control group [weighted mean difference (WMD) = -1.01, 95% confidence interval (CI) (-3.6 and 1.6), and p = 0.003], necrotic core (NC) volume [WMD =-5.41, 95% CI(-13.3 and 2.5), and p = 0.35], and change dense calcification (change DC) volume [WMD =-1.14, 95% CI (- 1.4 and – 0.8), and p = 0.62] between the treatment group and the control group.

**Conclusions:** Our study indicates that the addition of ezetimibe to statin therapy results in a modest yet significant further reduction in LDL-C compared to statin monotherapy. Ezetimibe led to a significant reduction in FFP volume; however, there were no statistically significant differences observed for NC, or change DC volumes.

## Introduction

Atherosclerotic cardiovascular disease (ACD) is the leading cause of mortality worldwide. ACD is primarily caused by elevated concentrations of low-density lipoprotein cholesterol (LDL-C) [1, 2]. To prevent future cardiovascular events, it is highly recommended to focus on the development of novel drugs that target and effectively lower lipoprotein levels. The use of tolerated statins is recommended for reducing LDL-C in patients with ACD. However, it is important to note that even with statin therapy, additional lipid-lowering therapies may be necessary for many patients with clinical ACD. Ezetimibe, a new cholesterol absorption inhibitor drug, has demonstrated the ability to further lower LDL-C levels [3–5]. In a randomized controlled trial, it was demonstrated that the combination of ezetimibe with statins significantly reduces levels of LDL-C [6]. Additionally, the combination of ezetimibe and statins leads to a substantial decrease in coronary plaque volume compared to statin treatment alone [7]. Although the effectiveness of ezetimibe in treating ACD has been acknowledged, there is an absence of a comprehensive meta-analysis regarding the efficacy of ezetimibe in LDL-C lowering in ACD patients. Hence, the objective of this study was to conduct a systematic evaluation of the effectiveness of ezetimibe in LDL-C lowering in ACD patients, to offer valuable insights for therapeutic approaches.

## Methods

### Search strategy

We conducted a comprehensive search in Pubmed/MEDLINE, EMBASE, and Cochrane Library to identify relevant studies published until July 1, 2023, that reported on the efficacy and effectiveness of ezetimibe in patients with ACD. The search terms used were ezetimibe, coronary artery, atherosclerosis, and randomized controlled trial. Only studies published in English were included. This study adhered to the Preferred Reporting Items for Systematic Reviews and Meta-Analyses statement (PRISMA) for its design and reporting (Prospero pending ID: 449721) [8].

### Study Selection

The retrieved records from the database searches were combined, and duplicates were eliminated using EndNote X7 (Thomson Reuters, Toronto, ON, Canada). Two reviewers (MR and MJN) independently assessed the records based on their title/abstract and full text to exclude any that were not relevant to the objectives of the study.

The studies included in the analysis met the following criteria:

*Participants:* The patients included in the studies had a diagnosis of ACD, which included individuals with a history of myocardial infarction, stable or unstable angina, coronary or other arterial revascularization, stroke, transient ischemic attack, or peripheral arterial disease.

*Intervention:* The intervention being investigated was the use of ezetimibe therapy, either as a standalone treatment or in combination with other lipid-lowering therapies.

*Comparison:* patients who received standard treatment or in combination with other lipid-lowering therapies.

*Outcome:* The primary outcome was the average change observed in LDL-C levels when compared to the baseline measurements.

### Data extraction

Two reviewers (MR and MJN) designed a data extraction form and extracted data from all eligible studies, with differences being resolved by consensus. The following data were extracted: first author’s name; year of publication; study duration; type of study; country or countries where the study was conducted; number of patients with ACD; patient age; treatment protocols; demographics (i.e., age, sex, nationality); and treatment outcomes.

### Quality assessment

The quality assessment of the included studies was conducted by two reviewers (MR and PM) using Cochrane tool [9]. In case of any discrepancies, a third reviewer (MJN) was involved. This tool encompasses various domains, including random sequence generation, allocation concealment, blinding of participants and personnel, blinding of outcome assessors, completeness of outcome data, and other factors such as selective reporting and other potential biases. Each study was categorized as having a low risk of bias when no concerns regarding bias were identified, a high risk of bias when there were concerns about bias, or an unclear risk of bias when there was insufficient information available.

### Data analysis

The statistical analysis was performed using Comprehensive Meta-Analysis software, version 3.0 (Biostat Inc., Englewood, NJ, USA). The weighted mean difference (WMD) was used as the pooled statistic, with a corresponding 95% confidence interval (CI). The degree of heterogeneity among the studies was assessed using the I2 value and p-value. In cases where the statistical heterogeneity between the studies was low (I2 ≤ 50% or p ≥ 0.1), the fixed-effect model was utilized. Conversely, if a significant level of inter-study heterogeneity was observed (I2 > 50% or p < 0.1), the random-effects model was employed. Cochran’s Q test and the I2 statistic were used to assess between-study heterogeneity. To evaluate publication bias, Begg’s test was applied, where a P value of less than 0.05 was considered indicative of statistically significant publication bias.

## Results

Figure 1 displays the systematic review flow diagram. Through the systematic literature review, a total of 20 records meeting the eligibility criteria were identified. These records reported the primary outcome, either the change in LDL-C from baseline or the LDL-C level at the endpoint, or both. They provided sufficient independent information for the meta-analysis. The inclusion/exclusion justifications are summarized, and Table 1 lists the 20 studies included in the meta-analysis. The table presents details about the study design, duration, baseline demographic characteristics.

**Figure 1.**
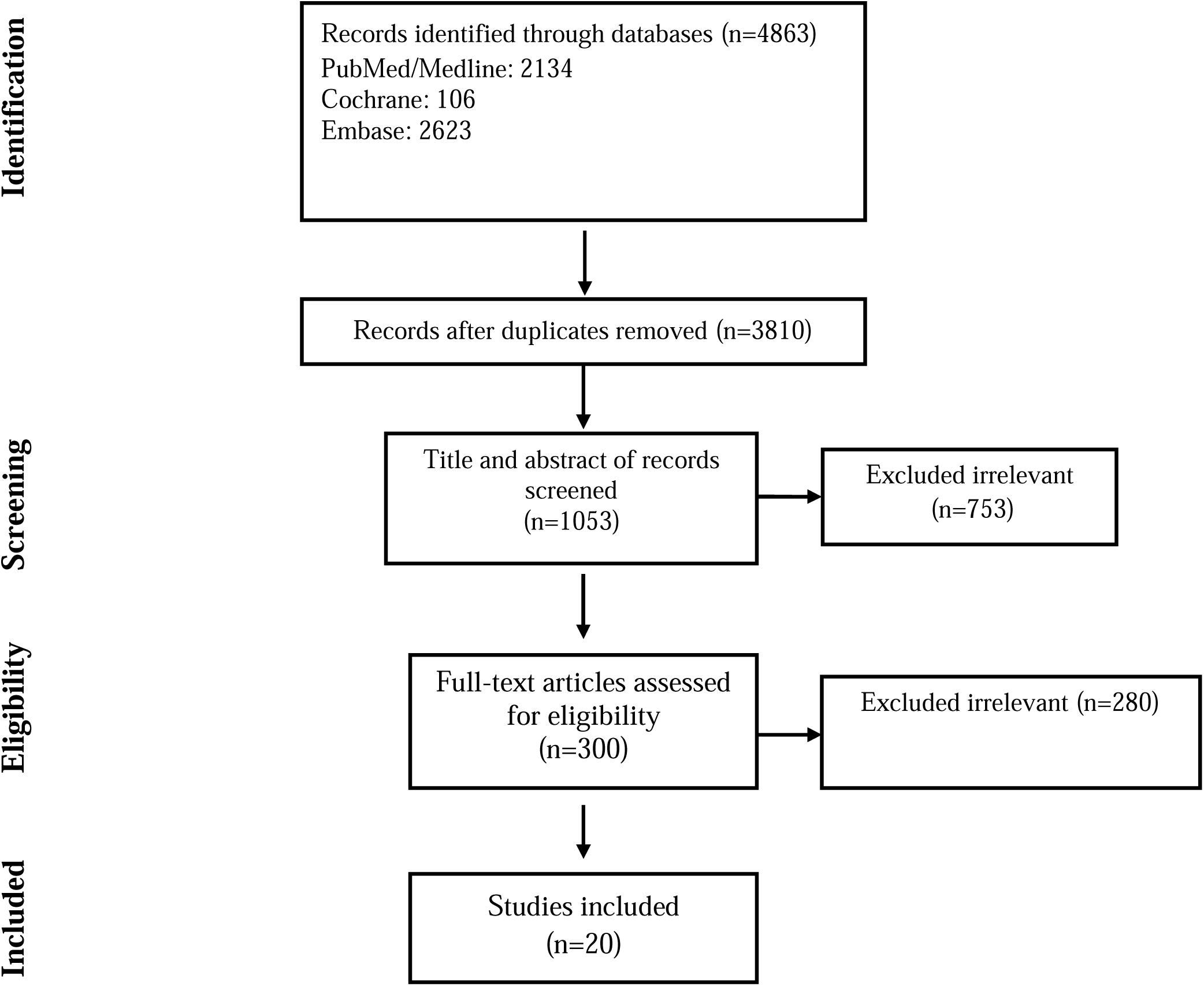
Flow chart of study selection for inclusion in the systematic review.

**Table 1.** Demographics and baseline characteristics of patients included in the meta-analysis according to study and treatment group.

| Authors | Study design | Sample size<br>T/C | Age<br>T | Intervention<br>T | C | Duration |
| --- | --- | --- | --- | --- | --- | --- |
| Hougaard,<br>2017 [13] | RCT | 43/44 | 55.3 ± 11.0 | EZ (10) + AT<br>(80) | PL (10) + AT<br>(80) | 12 months |
| Jung-(2016)<br>[14] | RCT | 34/36 | 60.9 ± 10.9 | EZ (10) + SI<br>(40) | PR (20) | 03 months |
| Kovarnik(201<br>2)<br>[15] | RCT | 42/47 | 63.5 ± 9.3 | EZ (10) + AT<br>(80) | AT (10) | 12 months |
| Brohet et al.<br>(2005)<br>[16] | RCT | 208/210 | 63.6± 11.1 | EZE 10 mg<br>QD+ SIM<br>10/20 mg QD | SIM 10/20<br>mg QD | 6 weeks |
| Cannon et al.<br>(2015)<br>[17] | RCT | 9067/9077 | 63.6± 9.7 | EZE 10 mg<br>QD + SIM<br>40–80 mg QD | IM 40–80 mg<br>QD | 6 years |
| Hibi et al.<br>(2018)<br>[18] | RCT | 50/53 | 63± 10.0 | EZE 10 mg<br>QD+PITA<br>2 mg | PITA 2 mg QD | 10 months |
| Joshi et al.<br>(2017)<br>[19] | RCT | 40/40 | 60.3± 9.8 | EZE 10 mg<br>QD +ROSU<br>10 mg QD | ROSU 10 mg<br>QD | 24 weeks |
| Masuda et al.<br>(2015)<br>[20] | RCT | 21/19 | 64.0± 7.9 | EZE 10 mg<br>QD + ROSU<br>5 mg QD | ROSU 5 mg<br>QD | 6 months |
| Ran et al.<br>(2017)<br>[21] | RCT | 42/42 | 60.4± 8.2 | EZE 10 mg<br>QD +ROSU<br>10 mg QD | ROSU 10 mg<br>QD | 10 weeks |
| Ren et al.<br>(2017)<br>[22] | RCT | 55/58 | 57.3± 1.5 | EZE 10 mg<br>QD +ROSU<br>10 mg QD | ROSU 10 mg<br>QD | 12 months |
| Ueda et al.<br>(2017)<br>[23] | RCT | 54/54 | 71± 8.0 | EZE 10 mg<br>QD + ATOR<br>10–20 mg QD | ATOR 10–20<br>mg QD | 9 months |
| Wang et al.<br>(2017)<br>[24] | RCT | 51/49 | 58± 10.0 | EZE 10 mg<br>QD +ATOR<br>20 mg<br>QD | ATOR 20 mg<br>QD | 12 months |
| Wang et al.<br>(2016)<br>[25] | RCT | 50/48 | 63± 10.0 | EZE 10 mg<br>QD +ROSU<br>10 mg QD | ROSU 10 mg<br>QD | 12 months |
| West et al.<br>(2011)<br>[26] | RCT | 18/33 | 62± 8.0 | EZE 10 mg<br>QD+SIM<br>40 mg QD | EZE 10 mg<br>QD | 2 years |
| Zou et al.<br>(2016)<br>[27] | RCT | 40/40 | 69.3± 5.8 | EZE 10 mg<br>QD +ATOR<br>10 mg QD | ATOR 10 mg<br>QD | 12 months |
| El-Tamalawy<br>et al (2018)<br>[28] | RCT | 33/32 | 61 ± 7.1 | EZE 10<br>mg/day +<br>ATOR 40 mg<br>daily | ATOR 80 mg<br>daily | 3 months |
| Klassen et<br>al(2021)<br>[29] | RCT | 10/10 | 62 (59–64) | EZE 10 mg<br>QD + ROSU<br>20 mg QD or<br>SIM 40 mg | ROSU 20 mg<br>QD | 1 month |
| Oh et<br>al(2021)<br>[30] | RCT | 18/19 | 56.3 § 7.1 | ATO10 mg +<br>EZE10 mg | ATO 40 mg | 12 months |
| Pinto et al(2021) [31] | RCT | 50/51 | 59 (52–65) | EZE 10mg+SIM 40 mg | ROSU 20 mg | 1 month |
| Blom et al(2014) [32] | RCT | 63/73 | 55.9±9.0 | EZE 10 mg+ ATOR 80 mg | ATOR 80 mg | 1-2 months |

T, treatment group; C, control group; EZ, ezetimibe; ATOR, atorvastatin; SIM, simvastatin

Although the objective was to evaluate LDL-C lowering with ezetimibe therapy, with or without other LLTs, all the included studies compared combination ezetimibe plus statin therapy with statin monotherapy. The mean age of participants across the studies ranged from 57 to 71 years, depending on the treatment group (Table 1).

### Quality assessment

Regarding the risk of bias across the 20 trials, insufficient detail was provided about the randomization methodology, resulting in an unclear risk of selection bias for random sequence generation and allocation concealment. Other biases were assessed as low in the trials (Table 2).

**Table 2.** Quality assessment.

| Author | Random sequence generation | Allocation concealment | Blinding of participants and personnel | Blinding of outcome assessment | Incomplete outcome data |
| --- | --- | --- | --- | --- | --- |
| Brohet et al | L | U | L | U | L |
| Cannon et al | U | U | U | U | L |
| Hibi et al | L | U | H | L | L |
| Joshi et al | L | U | U | U | L |
| Masuda et al | L | L | H | H | L |
| Ran et al | L | U | H | U | L |
| Ren et al | L | U | U | U | L |
| Ueda et al | L | U | H | L | L |
| Wang 2017 et al | U | U | U | U | L |
| Wang 2016 et al | L | U | U | U | L |
| West et al | L | U | L | L | L |
| Zou et al | U | U | U | U | L |
| El-Tamalawy et al | L | U | L | L | L |
| Klassen et al | L | U | L | L | L |
| Oh et al | L | U | L | L | L |
| Zou et al | U | U | U | U | L |
| El-Tamalawy et al | L | U | L | L | L |
| Klassen et al | L | U | L | L | L |
| Oh et al | L | U | L | L | L |
| Pinto et al | L | U | L | L | L |
| Blom et al | L | U | L | L | L |
| Mikkle et al | L | U | L | L | L |
| Jung et al | L | U | L | L | L |
| Kovarnik et al | L | H | L | L | L |

L; low, H; high, U; unclear

### Efficacy of LDL-C Lowering of Ezetimibe

As indicated in Figure 1, patients receiving combination of statin and ezetimibe were found to have a significant additional reduction in LDL-C (−14.06 mg/dL; 95% confidence interval [CI] −18.0 to −10.0, *p* < 0.0001) than those receiving statin alone.

**Figure 1.**
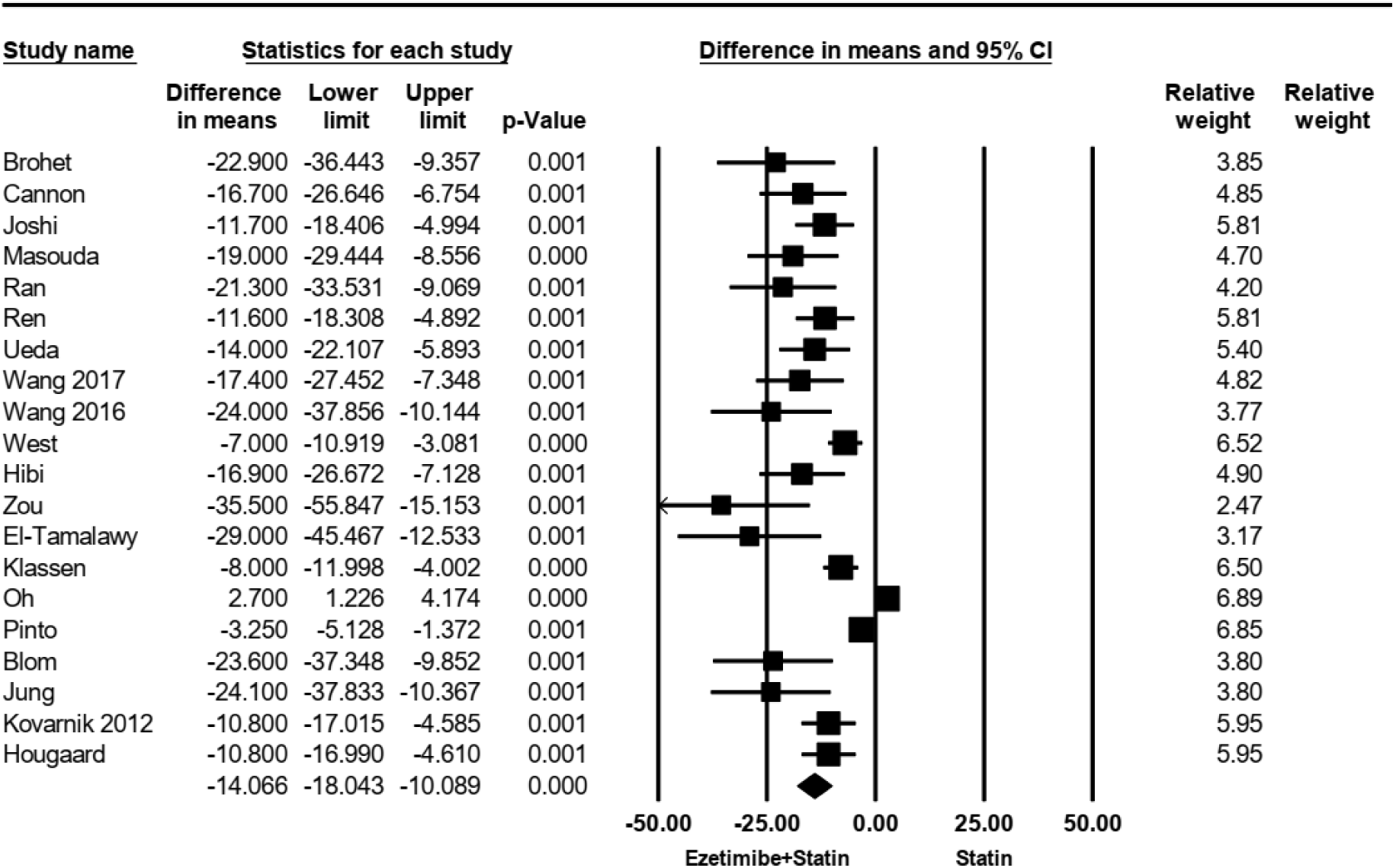
Treatment difference in mean LDL-C change (mg/dL) from baseline between combination ezetimibe plus statin therapy and statin monotherapy comparator at 6 months or at the reported timepoint closest to 6 months.

### Fibro-fatty plaque (FFP) volume

The efficacy of FFP was evaluated in four studies. There was no heterogeneity among the studies (I2 = 58.4%, p = 0.065). Using a random-effects model analysis, it was found that treatment interventions in the study group effectively reduced FFP when compared to the control group, showing a statistically significant difference [WMD = -1.01, 95% CI (-3.6 and 1.6), and p = 0.45], as shown in Figure2.

### Necrotic core (NC) volume

The efficacy of NC was reported in three out of the four research studies. There was no heterogeneity among the studies (I2 = 70.4%, p = 0.03). Using a random-effects model analysis, the results showed no significant difference in the reduction of NC between the treatment group and the control group [WMD = -5.41, 95% CI (-13.3 and 2.5), and p = 0.18], as shown in Figure 3.

**Figure 2.**
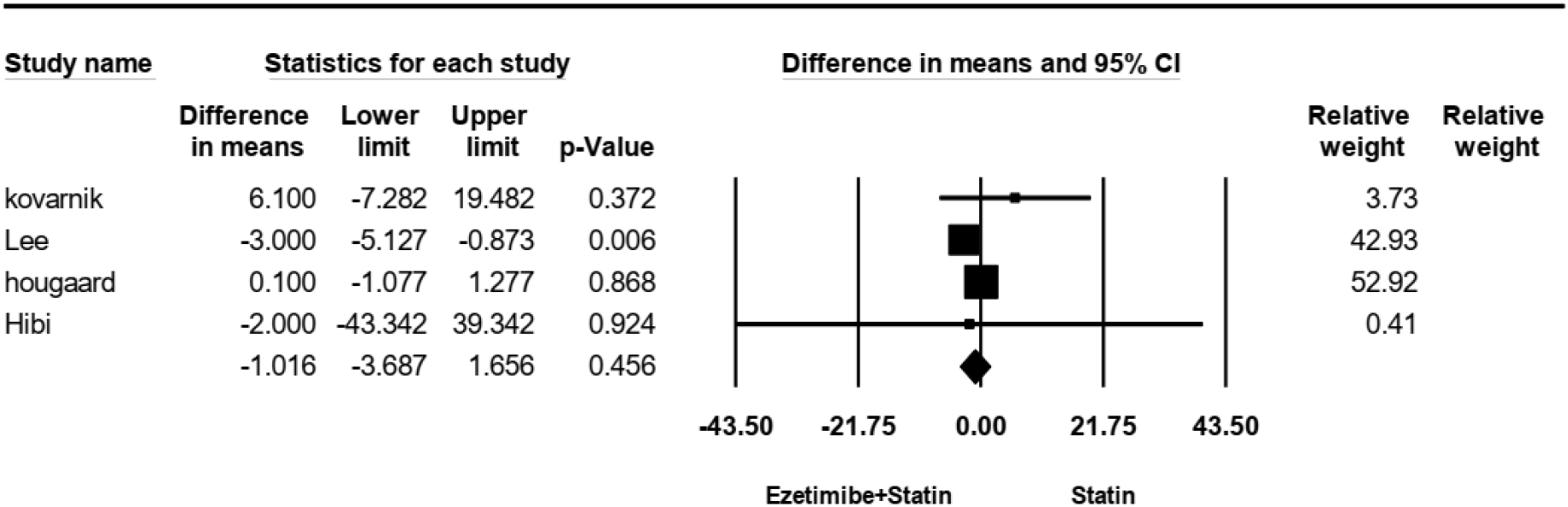
Forst plot for Fibro-Fatty plaque volume.

**Figure 3.**
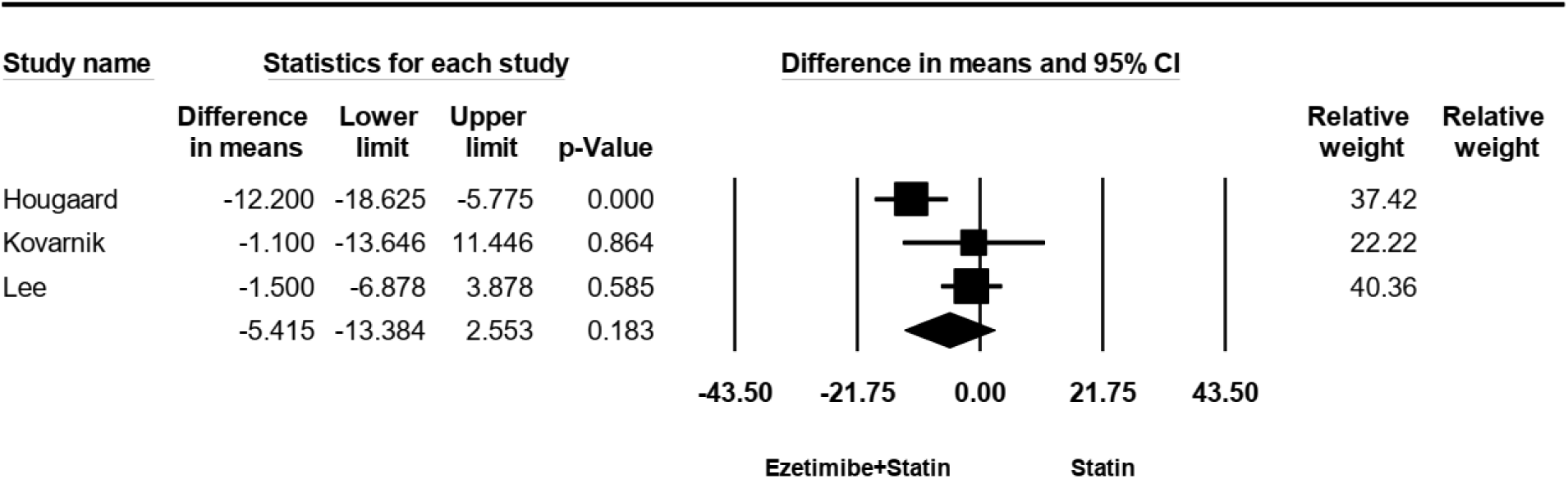
The forest plot for necrotic core volume.

### Change dense calcification (change DC) volume

All research studies examined the efficacy of change DC. There was no heterogeneity among the studies (I2 = 0%, p = 0.42). Using a fixed-effects model analysis, the results showed no significant difference in the reduction of change DC between the treatment group and the control group [WMD = -1.14, 95% CI (- 1.4and – 0.8), and p = 0.00], as shown in Figure 4.

**Figure 4.**
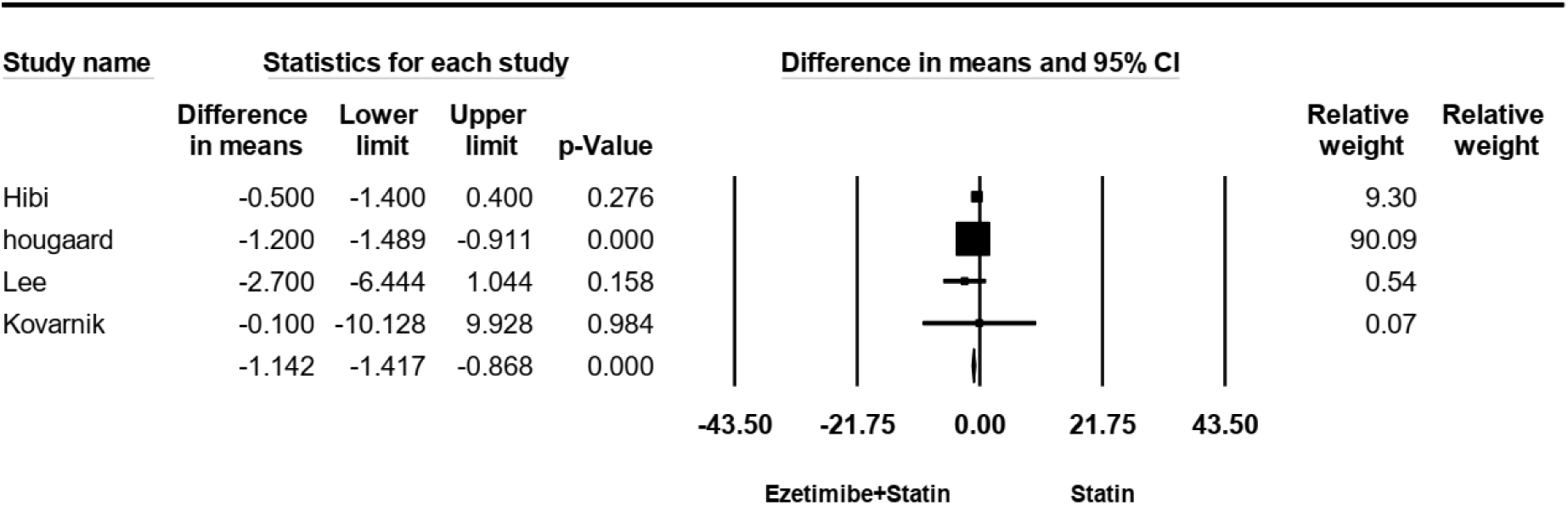
The forest plot for Dense calcification volume.

## Discussion

### Principal findings

In this meta-analysis we demonstrated that, for patients with ACD, ezetimibe added to statin therapy provided an additional reduction of LDL-C compared with statin monotherapy.

### Comparisons with other studies

Our results are consistent with prior evidence on lipid-lowering therapies. For instance, a meta-analysis by Shaya et al showed larger relative in LDL-C by addition of ezetimibe to statin therapy, compared with statin monotherapy [10]. Another meta-analysis demonstrated that ezetimibe significantly decreased coronary atherosclerotic plaque compared to the control group (placebo or statin monotherapy) [11]. Likewise, Toyota et al confirms that all 3 strategies for enhancing LDL-C reduction- i.e. more-intensive statin therapy, adding ezetimibe, and adding PCSK9, could improves clinical outcomes in high atherosclerotic cardiovascular disease [12].

### Strength and limitations of this study

The final result of our study is along with the Shaya et al. meta-analysis published in 2020 which demonstrated a decreased risk of ACD in patients receiving ezetimibe [10]. However, our study has some strengths and is different in some ways. We also included studies focused on the FFP volume, NC volume and DC volume. Studies in which the sample population had specific comorbidities were excluded to lessen the potential effect of the confounding factors. Also, we excluded the articles in which the full text was not found. Against this, we included eight other studies that not previously included. Our calculated WMD (−14.06, 95%CI 18.0-10.0) was a little higher than Shaya et al. (-21.8, CI 95% -26.5-,17.1) but in line with its direction and statistical significance.

The major limitation of our study is its originality, as the data on the efficacy of ezetimibe on LDL-C has been known and ascertained for some time and also reported by international guideline. However, an update of data relating to scientific evidence can still have value. Some other limitations of our study are associated with the studies we included and are as follows: First, there have not been any recent clinical trials studies on the topic and the most recent one was published in 2021. Further new investigations with a large number of populations are required to elucidate the issue. Second, there were confounding factors in the included studies which we have inability to subgroup analysis, due to insufficient and inconsistent data. Third, the mortality benefits of ezetimibe despite its intensive lowering of LDL-C levels should be investigated. The survival advantage relies on several factors, including the efficacy of the drug, competing risks, off-target effects, baseline cardiovascular risk, and follow-up duration of the study.

### Clinical implications

This systematic review informs decision makers about the benefits of ezetimibe on important cardiovascular outcomes. The key observations are that moderate to high certainty evidence favors ezetimibe for lowering LDL-C levels and. Furthermore, these agents lead to reducing the volume of FFP.

### Conclusions

Our study indicates that the addition of ezetimibe to statin therapy results in a modest yet significant further reduction in LDL-C compared to statin monotherapy. Ezetimibe led to a significant reduction in FFP volume; however, there were no statistically significant differences observed for NC, or change DC volumes.

## Data Availability

All data produced in the present work are contained in the manuscript

## References

1. Kronenberg, F., et al., Lipoprotein (a) in atherosclerotic cardiovascular disease and aortic stenosis: a European Atherosclerosis Society consensus statement. European heart journal, 2022. 43(39): p. 3925–3946.

2. Mortensen, M.B. and B.G. Nordestgaard, Elevated LDL cholesterol and increased risk of myocardial infarction and atherosclerotic cardiovascular disease in individuals aged 70-100 years: a contemporary primary prevention cohort. The Lancet, 2020. 396(10263): p. 1644–1652.

3. Ouchi, Y., et al., Ezetimibe lipid-lowering trial on prevention of atherosclerotic cardiovascular disease in 75 or older (ewtopia 75) A randomized, controlled trial. Circulation, 2019. 140(12): p. 992–1003.

4. Phan, B.A.P., T.D. Dayspring, and P.P. Toth, Ezetimibe therapy: mechanism of action and clinical update. Vascular health and risk management, 2012: p. 415–427.

5. Vavlukis, M. and A. Vavlukis, Adding ezetimibe to statin therapy: latest evidence and clinical implications. Drugs in Context, 2018. 7.

6. Murphy, S.A., et al., Reduction in total cardiovascular events with ezetimibe/simvastatin post-acute coronary syndrome: the IMPROVE-IT trial. Journal of the American College of Cardiology, 2016. 67(4): p. 353–361.

7. Ueda, Y., et al., Effect of Ezetimibe on Stabilization and Regression of Intracoronary Plaque―The ZIPANGU Study―. Circulation Journal, 2017. 81(11): p. 1611–1619.

8. Moher, D., et al., Preferred reporting items for systematic reviews and meta-analyses: the PRISMA statement. Annals of internal medicine, 2009. 151(4): p. 264–269.

9. Higgins, J.P., et al., The Cochrane Collaboration’s tool for assessing risk of bias in randomised trials. Bmj, 2011. 343.

10. Shaya, F.T., et al., Lipid-lowering efficacy of ezetimibe in patients with atherosclerotic cardiovascular disease: a systematic review and meta-analyses. American Journal of Cardiovascular Drugs, 2020. 20: p. 239–248.

11. Chai, B., et al., Meta-analysis and trial sequential analysis of ezetimibe for coronary atherosclerotic plaque compositions. Frontiers in Pharmacology, 2023. 14: p. 1166762.

12. Toyota, T., et al., More-Versus Less-Intensive Lipid-Lowering Therapy. Circ Cardiovasc Qual Outcomes, 2019. 12(8): p. e005460.

13. Hougaard, M., et al., Influence of ezetimibe in addition to high-dose atorvastatin therapy on plaque composition in patients with ST-segment elevation myocardial infarction assessed by serial: Intravascular ultrasound with iMap: the OCTIVUS trial. Cardiovasc Revasc Med, 2017. 18(2): p. 110–117.

14. Lee, J.H., et al., Early Effects of Intensive Lipid-Lowering Treatment on Plaque Characteristics Assessed by Virtual Histology Intravascular Ultrasound. Yonsei medical journal, 2016. 57(5): p. 1087–1094.

15. Kovarnik, T., et al., Virtual histology evaluation of atherosclerosis regression during atorvastatin and ezetimibe administration: HEAVEN study. Circ J, 2012. 76(1): p. 176–83.

16. Brohet, C., et al., LDL-C goal attainment with the addition of ezetimibe to ongoing simvastatin treatment in coronary heart disease patients with hypercholesterolemia. Curr Med Res Opin, 2005. 21(4): p. 571–8.

17. Cannon, C.P., et al., Ezetimibe added to statin therapy after acute coronary syndromes. New England Journal of Medicine, 2015. 372(25): p. 2387–2397.

18. Hibi, K., et al., Effects of Ezetimibe-Statin Combination Therapy on Coronary Atherosclerosis in Acute Coronary Syndrome. Circ J, 2018. 82(3): p. 757–766.

19. Joshi, S., et al., Efficacy of Combination Therapy of Rosuvastatin and Ezetimibe vs Rosuvastatin Monotherapy on Lipid Profile of Patients with Coronary Artery Disease. Journal of Clinical & Diagnostic Research, 2017. 11(12).

20. Masuda, J., et al., Effect of combination therapy of ezetimibe and rosuvastatin on regression of coronary atherosclerosis in patients with coronary artery disease. Int Heart J, 2015. 56(3): p. 278–85.

21. Ran, D., et al., A randomized, controlled comparison of different intensive lipid-lowering therapies in Chinese patients with non-ST-elevation acute coronary syndrome (NSTE-ACS): Ezetimibe and rosuvastatin versus high-dose rosuvastatin. Int J Cardiol, 2017. 235: p. 49–55.

22. Ren, Y., et al., Comparison of the effect of rosuvastatin versus rosuvastatin/ezetimibe on markers of inflammation in patients with acute myocardial infarction. Exp Ther Med, 2017. 14(5): p. 4942–4950.

23. Ueda, Y., et al., Effect of Ezetimibe on Stabilization and Regression of Intracoronary Plaque - The ZIPANGU Study. Circ J, 2017. 81(11): p. 1611–1619.

24. Wang, J., et al., Efficacy of ezetimibe combined with atorvastatin in the treatment of carotid artery plaque in patients with type 2 diabetes mellitus complicated with coronary heart disease. Int Angiol, 2017. 36(5): p. 467–473.

25. Wang, X., et al., Effects of Combination of Ezetimibe and Rosuvastatin on Coronary Artery Plaque in Patients with Coronary Heart Disease. Heart Lung Circ, 2016. 25(5): p. 459–65.

26. West, A.M., et al., The effect of ezetimibe on peripheral arterial atherosclerosis depends upon statin use at baseline. Atherosclerosis, 2011. 218(1): p. 156–62.

27. Zou, Y., et al. Effect of Ezetimibe Combined with Low-dose Atorvastain Calcium on Carotid Atherosclerosis in Elderly Patients with Coronary Heart Disease. in Journal of the American Geriatrics Society. 2016. WILEY-BLACKWELL 111 RIVER ST, HOBOKEN 07030-5774, NJ USA.

28. El-Tamalawy, M.M., et al., Effect of Combination Therapy of Ezetimibe and Atorvastatin on Remnant Lipoprotein Versus Double Atorvastatin Dose in Egyptian Diabetic Patients. J Clin Pharmacol, 2018. 58(1): p. 34–41.

29. Klassen, A., et al., Evaluation of two highly effective lipid-lowering therapies in subjects with acute myocardial infarction. Sci Rep, 2021. 11(1): p. 15973.

30. Oh, P.C., et al., Effect of Atorvastatin (10 mg) and Ezetimibe (10 mg) Combination Compared to Atorvastatin (40 mg) Alone on Coronary Atherosclerosis. Am J Cardiol, 2021. 154: p. 22–28.

31. Pinto, L.C.S., et al., Main differences between two highly effective lipid-lowering therapies in subclasses of lipoproteins in patients with acute myocardial infarction. Lipids Health Dis, 2021. 20(1): p. 124.

32. Blom, D.J., et al., A 52-week placebo-controlled trial of evolocumab in hyperlipidemia. N Engl J Med, 2014. 370(19): p. 1809–19.

